# Hidden in the cortical folds: Sex-specific sulcal pits patterns in schizophrenia

**DOI:** 10.1101/2025.09.29.25336863

**Authors:** Noemí Hostalet, Daniel Herrera-Escartín, Lluís Cobos-Aumatell, Mateo Berrío-Soto, Banu Ahtam, Amalia Guerrero-Pedraza, Pedro Canut, Jesús J Gomar, Pilar Salgado-Pineda, Josep Salavert, Nuria Ramiro, Emilio J Inarejos Clemente, Salvador Sarró, Raymond Salvador, Edith Pomarol-Clotet, Neus Martínez-Abadias, Kiho Im, Mar Fatjó-Vilas

**Affiliations:** FIDMAG Germanes Hospitalàries Research Foundation, Barcelona, Spain; Departament de Biologia Evolutiva, Ecologia i Ciències Ambientals (BEECA), Facultat de Biologia, Universitat de Barcelona (UB), Spain; Centro de Investigación Biomédica en Red de Salud Mental (CIBERSAM), Instituto de Salud Carlos III, Spain; Programa de Doctorat en Biomedicina, Universitat de Barcelona (UB), Spain; Freelance computing assessor, Barcelona, Spain; Environment and Society Research Group (MASO) – Faculty of Social and Human Sciences, University of Antioquia, Colombia; Fetal Neonatal Neuroimaging and Developmental Science Center, Division of Newborn Medicine, Boston Children’s Hospital, Boston, MA, USA; Department of Pediatrics, Harvard Medical School, Boston, MA, USA; Fundación Hospitalarias Sant Boi, Sant Boi de Llobregat, Barcelona, Spain; The Litwin-Zucker Alzheimer’s Research Center, The Feinstein Institutes for Medical Research, Manhasset, NY 11030, USA; Fundación Hospitalarias Barcelona, Barcelona, Spain; Servei de Radiologia, Hospital Sant Joan de Déu, Barcelona

**Keywords:** sulcal pits, schizophrenia, neurodevelopment, transcriptional profiles

## Abstract

**Background:** Prenatal brain development represents a vulnerability window for schizophrenia (SZ). To explore early alterations related to the disorder, we examined sulcal pits —lifespan stable cortical landmarks of fetal cortical folding— as markers of neurodevelopmental differences between SZ patients and healthy controls (HC).

**Methods:** T1-weighted MRI (1.5T) scans were obtained from 426 individuals (237 SZ patients: 173 males, 64 females; 189 HC: 93 males, 96 females). Sulcal pits were identified from white matter cortical surfaces reconstructed using FreeSurfer. Graphs were constructed for each lobe and hemisphere and compared based on different sulcal pits (SP) features: SP-Position, SP-Depth, SP-SurfaceAreaBasin, SP-Topology, and all combined features (SP-Combined). We tested: i) group differences in the sulcal pits heterogeneity between patients and HC; ii) the sulcal pits divergence in patients relative to HC; iii) the association between sulcal pots divergence and Positive and Negative Symptoms Scale (PANSS) scores; iv) the regional covariation between the sulcal pits heterogeneity differences and transcriptional profiles across lobes using Partial Least Squared regressions (PLS-R). All analyses were performed in the sex-pooled sample and stratified by sex.

**Results:** Patients showed increased heterogeneity compared to HC across hemispheres, frontal, temporal, and parietal lobes. Divergence analyses revealed reduced similarity with respect to HC, in the left frontal SP-Combined in males (negatively correlated with PANSS scores) and right parietal SP-Topology in females. PLS-R showed sex-specific molecular underpinnings related to the identified differences in the heterogeneity of sulcal pits.

**Conclusion:** This study provides novel evidence supporting sulcal pits as early neurodevelopmental markers of structural sex-specific brain variability in SZ.

## 1. INTRODUCTION

Well-established evidence highlights prenatal brain development as a critical vulnerability window, during which the complex interplay of genetic and environmental factors may increase the risk for neurodevelopmental disorders such as schizophrenia (SZ) (Birnbaum & Weinberger, 2024).

Regarding genetic contributors, several SZ-associated loci exhibit peak expression in the brain during mid-gestation (Birnbaum & Weinberger, 2017; Wen et al., 2024), while others demonstrate sex-specific expression patterns in the placenta (Ursini et al., 2023). These coordinated expression profiles underscore the influence of genetic risk factors in shaping susceptibility to the disorder.

Among environmental factors, obstetric complications, as markers of prenatal stress, have not only been related to SZ (Mezquida et al., 2018) but also to cognitive impairments detected at the onset of the disorder (Amoretti et al., 2022). Furthermore, when distinguishing obstetric complications into antenatal and perinatal, those associated with the disorder were primarily antenatal, highlighting the importance of the impact of early insults on fetal brain development (Valli et al., 2023).

This gene-environment interplay contributes to the specific adult brain phenotypes associated with SZ (Van Erp et al., 2016; van Erp et al., 2018). However, despite considerable progress, it remains challenging to discern whether the neuroanatomical differences observed in patients reflect disrupted neurodevelopment trajectories or emerge later as consequences of the disorder progression. While indirect prenatal neurodevelopment markers —such as dermatoglyphic anomalies, which form during gestation and remain stable throughout life—, have been linked to both genetic and environmental SZ risk (Fatjó-Vilas et al., 2008; Golembo-Smith et al., 2012; Guardiola-Ripoll et al., 2024), there is still a need for innovative approaches capable of detecting brain-based features more directly tied to the earliest stages of prenatal brain development. To address this gap, we propose a novel approach centered on analyzing sulcal pits patterns.

Sulcal pits, defined as the deepest points of the cortical surface, are located along the bottom of sulcal lines (Auzias et al., 2015; Hostalet, et al., 2025; Im et al., 2010; Im & Grant, 2019; Lohmann et al., 2008). They emerge with the earliest cortical folds during fetal development, continue to form throughout subsequent prenatal stages, and remain stable across the lifespan (Meng et al., 2014; Solhtalab et al., 2025). Sulcal pits are considered markers of neurodevelopmental processes as their patterns have been differentially associated with neurodevelopmental disorders such as autism (Brun et al., 2016), dyslexia (Im et al., 2016), attention deficit hyperactivity disorder (Li, Jiang, et al., 2021; Li, Wang, et al., 2021), and more recently, with specific symptomatology in SZ (Lerosier et al., 2024; Salgado-Pineda et al., 2025). Although previous studies have analyzed the quantitative patterns of sulcal pits in SZ (Lerosier et al., 2024; Salgado-Pineda et al., 2025), none have examined their global pattern by considering not only the number and depth of sulcal pits, but also their spatial arrangement and interconnectivity, as proposed by Im et al., (2011). This approach provides deeper insight into the holistic development and morphometry of the cortex, as it enables the examination of the overall sulcal pattern in 3D by constructing sulcal graphs, where sulcal pits represent the nodes and their connections constitute the edges. Therefore, graph-based analysis of sulcal pits patterns represents a promising method for identifying early neurodevelopmental brain markers in SZ.

In addition to their morphological significance, the heritability of sulcal pits has been estimated to range between 0.2 and 0.5 (Le Guen et al., 2018). However, no study to date has explored their genetic architecture. In this context, public resources such as the Allen Human Brain Atlas (Hawrylycz et al., 2012; Shen et al., 2012) provide the opportunity to spatially integrate diagnosis-specific brain phenotypes, such as sulcal pits patterns, with gene expression data, thereby enabling the identification of genetic pathways that hypothetically may underlie diagnosis-specific neurodevelopmental profiles.

The general aim of this study was to investigate differences in the patterns of sulcal pits between patients and controls and their association with clinical and brain transcriptomic profiles. Previous studies underscoring the high heterogeneity in brain morphometry among patients with SZ (Alnæs et al., 2019; Fang et al., 2022; Janssen et al., 2021; Wolfers et al., 2018) highlight the importance of not only comparing patient groups to healthy individuals in terms of mean differences but also examining measures of within-group heterogeneity.

Accordingly, the specific aims of the study were: i) to compare the divergence in the sulcal pits patterns of SZ patients and healthy controls relative to the average of the healthy control group; ii) to assess differences in the heterogeneity of sulcal pits patterns between patients and healthy individuals; iii) to examine the correlation between the degree of divergence in the SZ patterns of sulcal pits (relative to controls) and clinical symptoms; and iv) to explore the spatial covariation between the heterogeneity of sulcal pits in SZ patients and brain gene expression profiles derived from the healthy individuals included in the Allen Human Brain Atlas (Hawrylycz et al., 2012; Shen et al., 2012).

Considering the sex-specific trajectories of brain development (Díaz-Caneja et al., 2021) and the observed sex differences in sulcal pit depth (Hostalet, et al., 2025), along with differences between males and females in the onset (Hoeksema et al., 2025), symptomatology (Amoretti et al., 2024; Bergen et al., 2014) and SZ-associated neurodevelopmental markers (Fatjó-Vilas et al., 2008; Hostalet et al., 2024), we aimed to perform all analyses both in the sex-pooled sample and separately by sex, in order to identify sex-specific patterns of sulcal pits associated with the disorder and their underlying genetic mechanisms.

## 2. METHODS

### 2.1. Sample

The sample comprised 426 subjects, including 237 patients with SZ and 189 healthy controls (HC) (Table 1). Patients were recruited at Fundació Hospitalàries Sant Boi and Fundació Hospitalàries Barcelona. According to a clinical interview with psychiatrists, they fulfilled the Diagnostic and Statistical Manual of Mental Disorders (4th edition, text revision) (DSM-IV-TR) criteria for SZ. HC consisted of non-medical staff, their relatives and acquaintances, plus public advertising in the area of Barcelona. They were questioned and excluded if they reported a personal history of mental illness and/or treatment with psychotropic medication or a first-degree family history of psychiatric disorders. In addition, both patients and HC met additional inclusion criteria, including European ancestry; age between 18 and 65 years old; right-handedness; and estimated intelligence quotient (IQ) > 70, as assessed using the *Test de Acentuación de Palabras* (TAP) (Gomar et al., 2011), an adapted Spanish version of the National Adult Reading Test (NART) (Nelson & Willison, 1991). Exclusion criteria for both groups included drug or alcohol abuse and a history of neurological damage.

**Table 1.** Sample characteristics of the individuals included in the study. The description is provided for the overall sample as well as separately for males and females.

|  | n | Mean age (SD) | Age group differences | PANSS<sup>a</sup> | PANSS-P | PANSS-N | PANSS-GP |
| --- | --- | --- | --- | --- | --- | --- | --- |
| HC | 189 | 37.58 (10.7) | t =-1.85;<br>p = 0.06 | - | - | - | - |
| SZ patients | 237 | 39.66 (11.8) |  | 74.59 (17.95) | 17.31 (5.81) | 21.81 (6.93) | 35.47 (9.05) |
| Male HC | 93 | 36.62 (10.70) | t =-1.58;<br>p = 0.12 | - | - | - | - |
| Male SZ patients | 173 | 38.95 (11.88) |  | 76.08 (18.1) | 17.46 (5.96) | 22.48 (6.72) | 36.14 (9.25) |
| Female HC | 96 | 38.51 (11.73) | t =-1.65;<br>p = 0.10 | - | - | - | - |
| Female SZ patients | 64 | 41.57 (11.08) |  | 70.26 (16.93) | 16.89 (5.39) | 19.85 (7.21) | 33.53 (8.25) |
| PANSS differences between male SZ and female SZ patients |  |  |  | t = 2.12;<br>p = 0.04 | t = 0.62;<br>p = 0.54 | t = 2.41;<br>p = 0.02 | t = 1.82;<br>p = 0.07 |

<sup>a</sup> PANSS assessment was available for 87% of the sex-pooled patients (88% of male patients and 83% of female patients)

### 2.2. Clinical assessment

Symptom severity in patients was evaluated with the Positive and Negative Symptoms Scale (PANSS) (Kay et al., 1987), administered by trained psychiatrists. The scores for the overall sample of patients and separately for males and females in the three subscales, positive symptoms (PANSS-P), negative symptoms (PANSS-N), general psychopathology (PANSS-GP), as well as the total score (PANSS), are detailed in Table 1. Differences between males and females with SZ were found for PANSS and PANSS-N (Table 1).

### 2.3. MRI data acquisition

High-resolution T1 weighted (T1w) head magnetic-resonance images (MRI) were obtained for all participants. The neuroimaging protocol was conducted in a 1.5T Signa Excite GE Medical Systems scanner at Hospital Sant Joan de Déu (Esplugues de Llobregat, Spain) using a 3D spoiled gradient echo sequence (SPGR) with the following acquisition parameters: (TR) = 12.4 ms, echo time (TE) = 5.2 ms, inversion time (TI) = 450 ms, flip angle = 20°, field of view (FOV) = 240 mm, acquisition matrix = 256 × 224, and reconstruction matrix = 512 × 512. Slice thickness was 2 mm with 180 slices acquired, resulting in an isotropic voxel size of approximately 0.47 × 0.47 × 2.0 mm. The pixel bandwidth was 61.05 Hz/pixel.

### 2.4. Image processing and sulcal pattern analysis

T1w MRIs were processed through the recon-all pipeline of FreeSurfer (v. 5.3), which included motion correction, removal of non-brain tissue, automated Talairach transformation, tessellation of the grey and white matter boundaries and topological correction to rectify holes, intersections and isolated vertices. The triangulated mesh representing the outer cortical surface was achieved using a deformation algorithm. All the images included in this study complied with the standardized quality control protocols from the ENIGMA consortium (http://enigma.ini.usc.edu/protocols/imaging-protocols).

Sulcal depth maps on the white matter surface were generated using FreeSurfer v5.3.0, and sulcal pits and their sulcal basins were automatically identified based on the smoothed sulcal depth map using a watershed segmentation algorithm (Im et al., 2010).

For sulcal pattern analysis, the global sulcal pattern was represented with a graph structure for each hemisphere and lobe using the sulcal pits as nodes and the connections of those sulcal pits placed in adjacent sulcal catchment basins as edges (Figure 1A).

**Figure 1.**
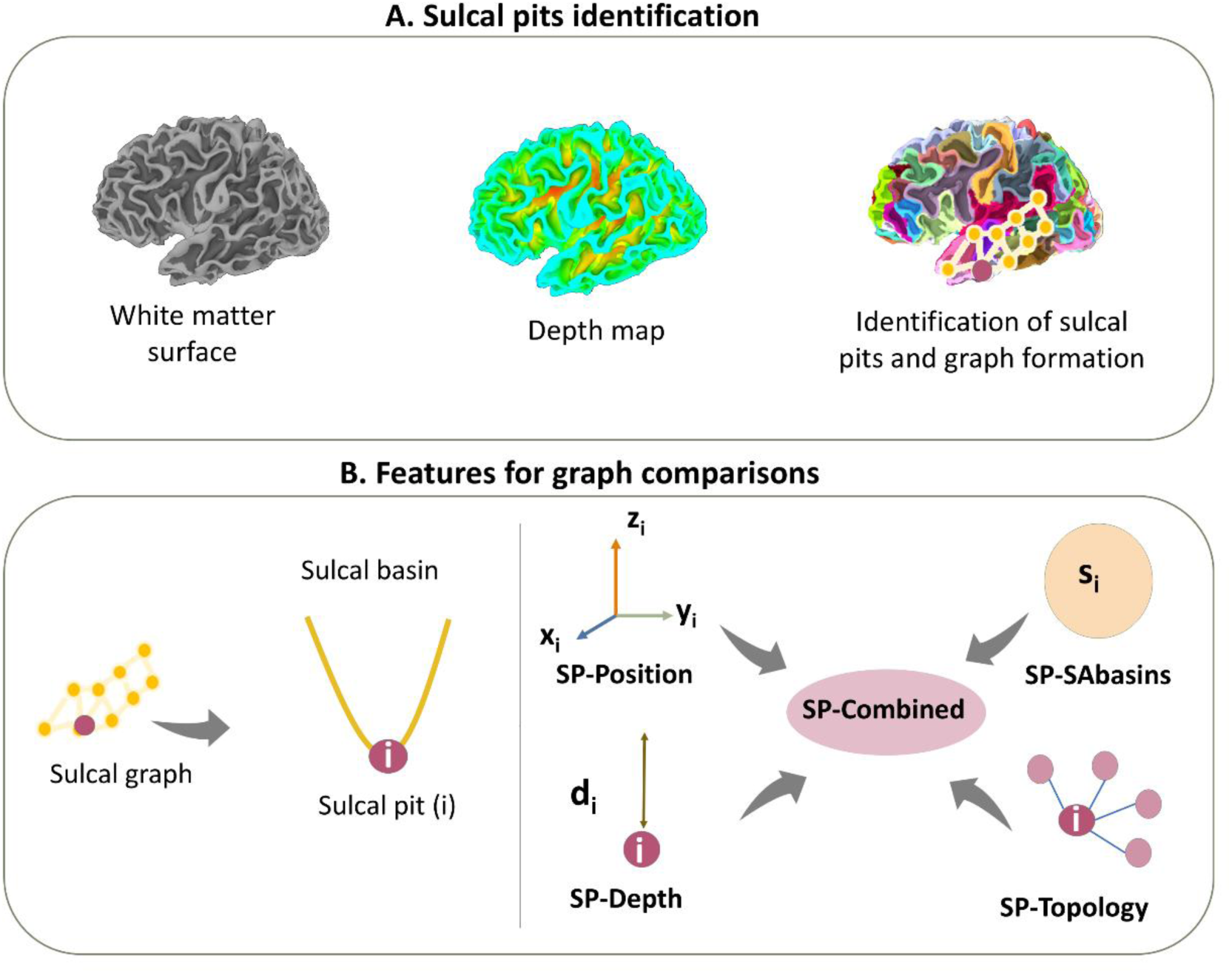
Representation of sulcal pits identification (A) and the graph features used for group comparison (B).

To characterize the global sulcal pattern, the graphs integrated the following features of sulcal folding: the 3D position (SP-Position), the depth of sulcal pits (SP-Depth), the surface area of sulcal basins (SP-SAbasins) and the graph topology (SP-Topology), a measure that included the number of edges and paths connecting the nodes (Figure 1B).

Each individual’s sulcal graphs were automatically compared using a spectral-based sulcal pattern matching and comparison technique for each hemisphere and lobe (Im et al., 2011), yielding three types of similarity values: within HC, within SZ patients and between HC – SZ patients for each feature (SP-Position, SP-Depth, SP-SAbasins, and SP-Topology) and all features combined (SP-Combined). The implemented statistical comparisons are presented in Figure 2. Specifically, the averaged similarity values were computed as follows: the within HC the within HC by averaging mean similarities between all possible pairs of HC individuals (Figure 2A); the within SZ by averaging mean similarities between all possible pairs of patients (Figure 2B); and the between SZ and HC by averaging mean similarities between each SZ patient and all HC (Figure 2C). The sulcal pattern similarity values ranged from 0 (non-existent similarity) to 1 (maximum similarity) (Im et al., 2011). These methodological procedures are explained in more detail in previous studies (Ahtam et al., 2021; Im et al., 2011, 2013; Morton et al., 2020, 2021).

**Figure 2.**
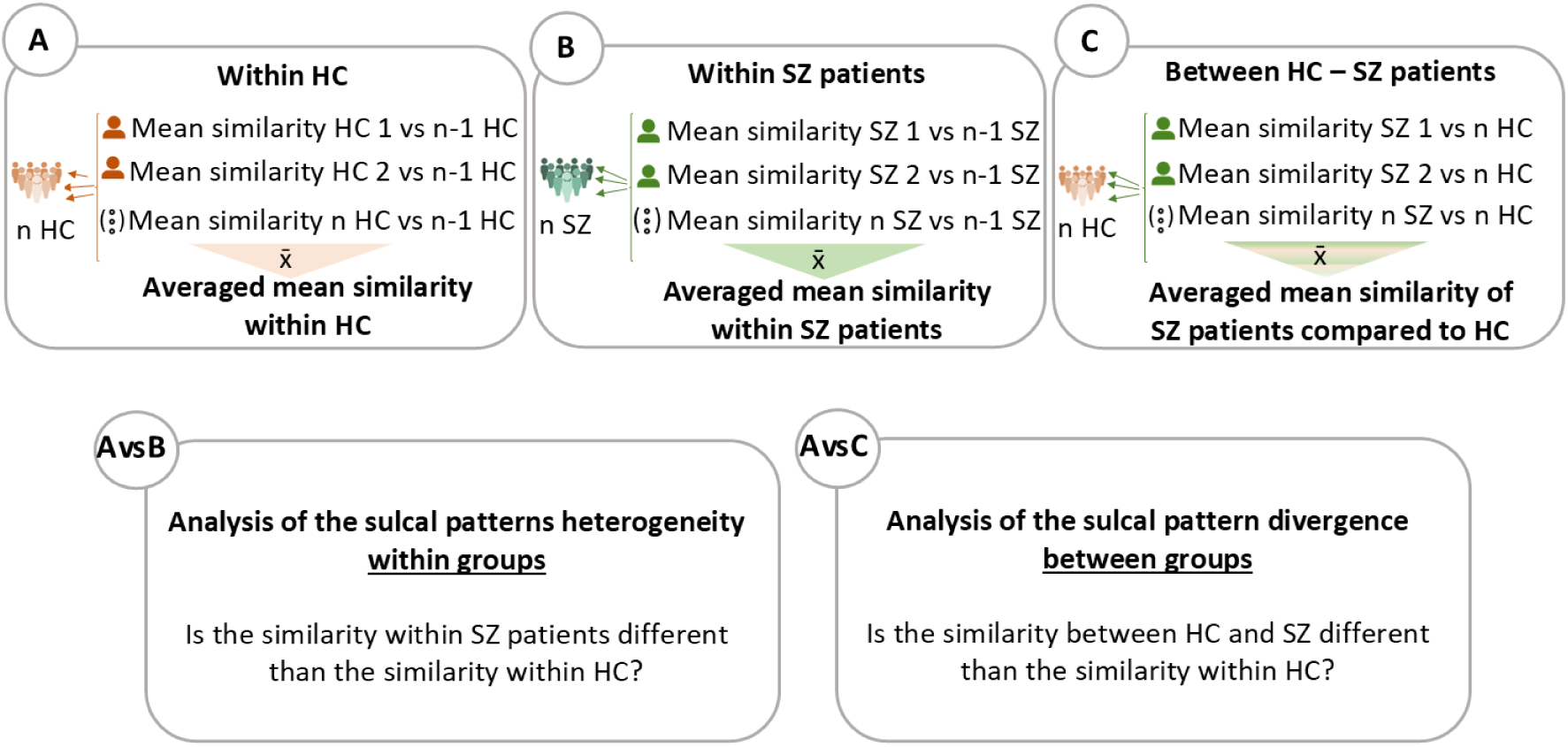
Schematic representation of the statistical comparisons. To obtain the averaged mean similarity within HC (A) the following steps were conducted: 1) each control was compared to each other control (obtaining n-1 values), 2) with the obtained values a mean similarity score for each control individual was computed, 3) the mean similarities of all controls were averaged, resulting in the averaged mean similarity score within HC. To obtain the averaged mean similarity within SZ patients (B), the same steps as for (A) were applied to SZ patients. Similarly, to obtain the averaged mean similarity between HC – SZ (C), each SZ patient was compared to each HC and the resulting mean similarities of patients were averaged. Then, to test differences in the heterogeneity of the sulcal patterns between groups, the averaged mean similarity within HC vs the averaged mean similarity within SZ patients (A vs B) were compared. Also, to assess the extent to which the sulcal patterns of patients diverged from those of HC, the averaged mean similarity within HC was tested against the averaged mean similarity of SZ patients compared to HC (A vs C).

To examine differences in the sulcal pattern heterogeneity within groups, the mean similarity within HC (Figure 2, A) was compared to the mean similarity within SZ patients (Figure 2, B). To evaluate the extent to which sulcal patterns in individuals with SZ diverge from those of HCs, the mean similarity within HC (Figure 2, A) was compared to the mean similarity between HC - SZ patients (Figure 2, C).

Group comparisons of mean similarity were conducted using t-tests. First, t-tests were applied to the raw similarity values. Second, to account for potential age effects, a linear regression was performed with age as a covariate on the raw similarity values, and the resulting residuals were used for subsequent t-tests. The resulting p-values were adjusted for multiple comparisons using the False Discovery Rate (pFDR) method (Benjamini & Hochberg, 1995).

### 2.5. Association between sulcal patterns and schizophrenia symptoms

In the group of patients, for those lobes where a statistically significant difference for any sulcal pits feature was detected in the between groups analysis (A vs C), we tested the associations of these features with the PANSS scores (PANSS total, PANSS-P, PANSS-N and PANSS-GP) through correlation analyses. This allowed assessing whether the degree of divergence from HC sulcal patterns correlated with the severity of SZ symptoms. Analyses were conducted in the sex-pooled sample and separately by sex and using both age-corrected and age-uncorrected similarity values.

### 2.6. Transcription-neuroimaging association analysis

The expression of 2,511 genes (Supplementary Materials, Table S1) with a consistent expression profile in the human cerebral cortex was obtained for each of the 34 hemispheric cortical regions from the Desikan Killiany atlas using data from the Allen Human Brain Atlas (six donors: five males and one female) (Hawrylycz et al., 2012). The selected genes exhibited stable inter-regional expression profiles across two independent postmortem datasets: the Allen Human Brain Atlas and the BrainSpan atlas, as defined in Shin et al. (2018). This procedure and its validity have been described in detail by French & Paus (2015) and Shin et al. (2018). Gene expressions of the Desikan-Killiany regions contained in each lobe (Klein & Tourville, 2012) were averaged to obtain expression values for each lobe and the whole hemisphere (Supplementary Materials, Table S1). To investigate the covariation between gene expression and the differences in sulcal pattern heterogeneity observed between HC and patients with SZ across brain regions, we conducted Partial Least Squares Regression (PLS-R) analyses separately for males and females, and following the guidelines in Han et al., (2025); Rohart et al. (2017) and Wold et al., (2001).

In the design of the PLS-R models, the response matrix (Y) comprised the age-corrected differences in the heterogeneity within groups for each lobe and hemisphere (age-corrected Cohen’s d values from the A vs. B comparison), while the predictor matrix (X) included the gene expression levels in each corresponding region. PLS-R models were designed with 2 components and their performances were assessed using repeated M-fold cross-validation (5 folds, repeated 10 times), yielding a Q^2^ estimate for each component. Values greater than 0 indicate overall good model performance. To provide a per-variable estimate of performance, the ratio Predictive Residual Sum of Squares/Sum of squares (PRESS/SS) was calculated for each Y-variable in the first component. According to Wold et al. (2001), Y-variables with PRESS/SS < 0.90 were adequately predictable for the PLS-R model. To assess whether the observed model performance for each Y-variable was significantly better than chance, a permutation test was conducted on the PRESS/SS values. The empirical p-value for each outcome was then estimated as the proportion of permuted PRESS/SS values that were equal to or less than the observed PRESS/SS value from the real model. The proportion of variance explained by each component and Y-variable was also calculated (Y-R^2^).

To identify which gene expressions contributed most to the modelling of the covariance between X and Y, the Variable Importance in Projection (VIP) scores were calculated for each variable in the X matrix. VIP scores serve as a metric in multi-variate data interpretation that offers insights into the level to which a variable contributes towards explaining the variation in the response variables (Chong & Jun, 2005). Subsequently, the top 10% of genes with the highest VIP scores were selected for Gene Ontology (GO) enrichment analysis using the online tool PANTHER (https://pantherdb.org/) with the aim of generating hypotheses regarding the functions of the genes most strongly contributing to sulcal pattern differences.

## 3. RESULTS

### 3.1. Analysis of the sulcal pattern heterogeneity within groups

Regarding the comparison of sulcal pattern heterogeneity between groups (A vs B) in the sex-pooled sample, differences were found in the left whole hemisphere and the left frontal lobe; as well as in the whole right hemisphere and the right frontal, temporal and parietal lobes (Table 2, Figure 3A). In most of the regions and measures showing differences, patients presented more heterogeneous sulcal patterns than HC (i.e. lower within group mean similarity in patients versus HC), and such differences remained significant after correcting for age. Only in the case of SP-Position and SP-Depth of the right parietal lobe, patients showed an opposite pattern (i.e. higher within group mean similarity in patients versus HC), but these effects were no longer significant when correcting for age (Table 2).

**Figure 3.**
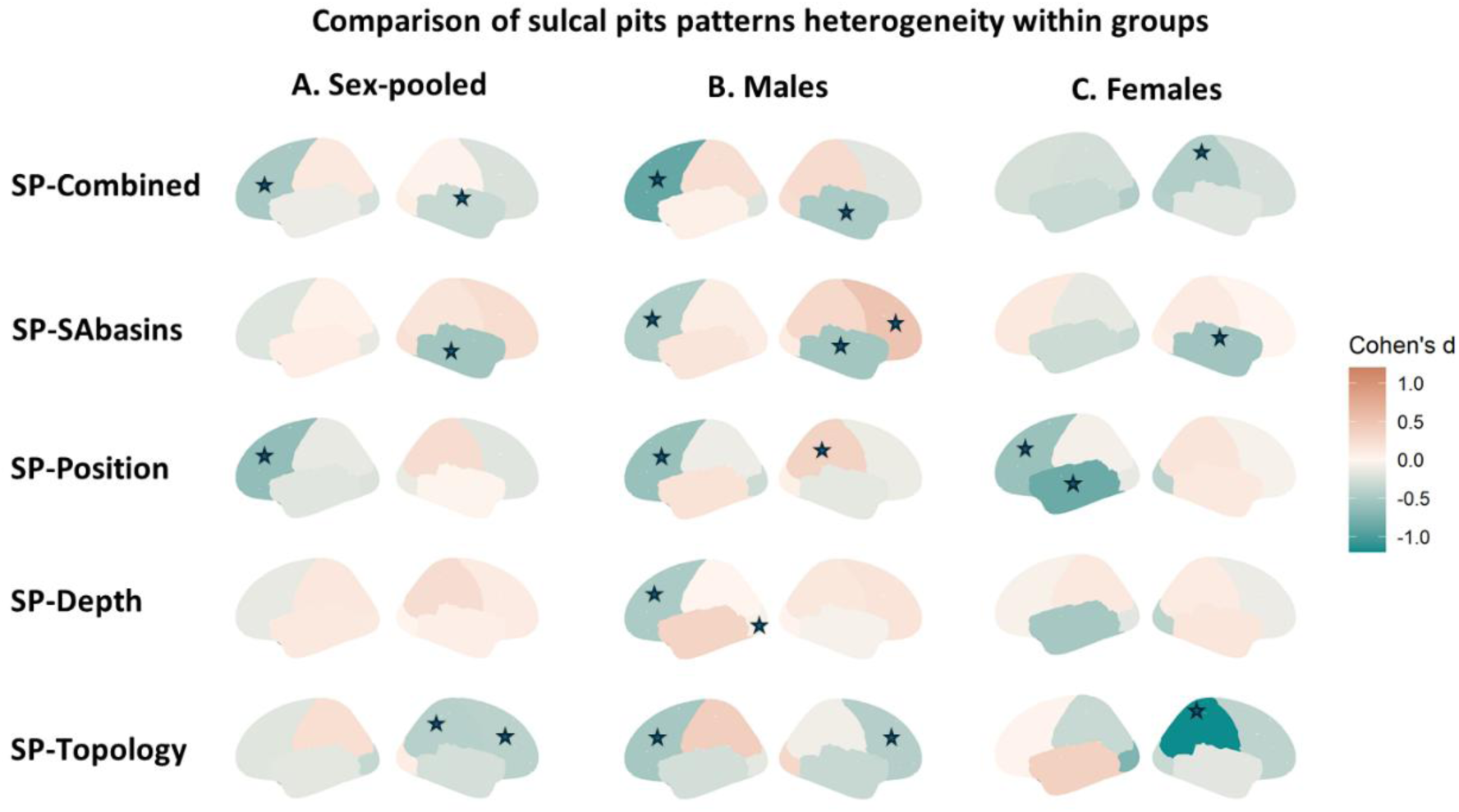
Within group differences in the mean similarity of the sulcal pits patterns. The figure is based on the effect sizes (Cohen’s d) of the age-corrected differences in sulcal pattern heterogeneity when comparing within SZ patients and within HCs across lobes in the sex-pooled (A), males (B) and females (C) samples. Stars indicate lobes showing age-corrected significant differences.

**Table 2.**
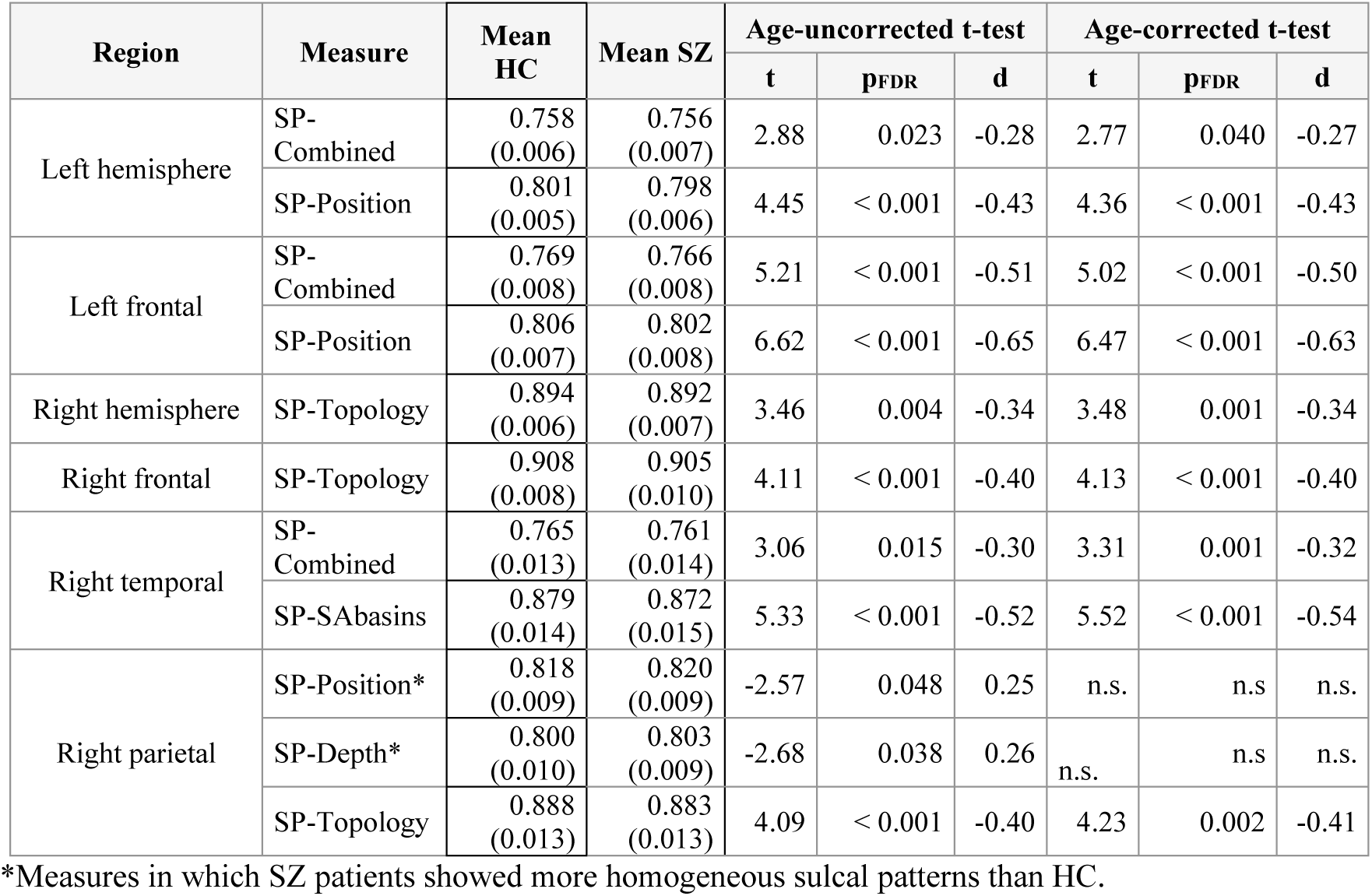
Brain regions and measures in which significant differences in the heterogeneity of the sulcal patterns within schizophrenia (SZ) patients and within HC were found in the sex-pooled sample. Results are shown before correcting for age (Age-uncorrected t-test) and after age correction (Age-corrected t-test).

Within males, differences were localized in the bilateral frontal, temporal and parietal lobes; and in the left occipital lobe (Table 3, Figure 3B). The predominant pattern was a greater heterogeneity in patients than in HC, except for the left temporal SP-Depth, left parietal SP-Topology, right frontal SP-SAbasins and right parietal SP-Position.

**Table 3.** Brain regions and sulcal pits pattern measures in which significant heterogeneity differences were detected between patients with schizophrenia (SZ) and healthy controls (HC) within the male sample. Results are shown before and after adjusting by age (Age-uncorrected t-test, Age-corrected t-test, respectively).

| Region | Measure | Mean<br>HC<br>(SD) | Mean SZ<br>(SD) | Age-uncorrected t-test |  |  | Age-corrected t-test |  |  |
| --- | --- | --- | --- | --- | --- | --- | --- | --- | --- |
|  |  |  |  | t | pFDR | d | t | pFDR | d |
| Left frontal | SP-Combined | 0.772<br>(0.008) | 0.766<br>(0.007) | 7.05 | <0.001 | -0.91 | 6.83 | < 0.001 | -0.88 |
|  | SP-Position | 0.806<br>(0.006) | 0.802<br>(0.008) | 4.71 | < 0.001 | -0.61 | 4.58 | < 0.001 | -0.59 |
|  | SP-Depth | 0.831<br>(0.007) | 0.828<br>(0.007) | 3.77 | 0.002 | -0.49 | 3.70 | 0.002 | -0.48 |
|  | SP-SAbasins | 0.900<br>(0.008) | 0.896<br>(0.007) | 3.69 | 0.002 | -0.48 | 3.51 | 0.003 | -0.45 |
|  | SP-Topology | 0.908<br>(0.008) | 0.903<br>(0.009) | 4.21 | < 0.001 | -0.54 | 4.00 | 0.001 | -0.51 |
| Left temporal | SP-Depth* | 0.820<br>(0.013) | 0.824<br>(0.013) | -2.82 | 0.020 | 0.36 | -2.60 | 0.040 | 0.33 |
| Left parietal | SP-Topology* | 0.882<br>(0.014) | 0.887<br>(0.014) | -3.07 | 0.011 | 0.40 | -3.04 | 0.010 | 0.39 |
| Left occipital | SP-Depth | 0.814<br>(0.012) | 0.808<br>(0.016) | 3.13 | 0.001 | -0.40 | 3.18 | 0.008 | -0.41 |
| Right frontal | SP-SAbasins* | 0.901<br>(0.006) | 0.904<br>(0.007) | -4.01 | 0.001 | 0.53 | -3.93 | 0.001 | 0.51 |
|  | SP-Topology | 0.909<br>(0.007) | 0.905<br>(0.001) | 3.28 | 0.007 | -0.42 | 3.37 | 0.005 | -0.43 |
| Right temporal | SP-Combined | 0.767<br>(0.013) | 0.761<br>(0.014) | 3.50 | 0.004 | -0.45 | 3.79 | 0.002 | -0.49 |
|  | SP-SAbasins | 0.879<br>(0.013) | 0.872<br>(0.015) | 3.88 | 0.001 | -0.50 | 4.11 | < 0.001 | -0.53 |
| Right parietal | SP-Position* | 0.817<br>(0.008) | 0.820<br>(0.009) | -2.87 | 0.0186 | 0.37 | -2.67 | 0.030 | 0.34 |

\*Measures in which SZ patients showed more homogeneous sulcal patterns than HC.

In females, differences were observed in the left hemisphere, the bilateral frontal and temporal lobes, and the right parietal lobe (Table 4, Figure 3C), with patients showing a greater heterogeneity than HC. Both in males and females, the differences remained significant when correcting for age.

**Table 4.** Brain regions and sulcal pits pattern measures in which significant heterogeneity differences were detected between patients with schizophrenia (SZ) and healthy controls (HC) within the female sample. Results are shown before and after adjusting by age (Age-uncorrected t-test, Age-corrected t-test, respectively).

| Region | Measure | Mean<br>HC<br>(SD) | Mean SZ<br>(SD) | Age-uncorrected t-test |  |  | Age-corrected t-test |  |  |
| --- | --- | --- | --- | --- | --- | --- | --- | --- | --- |
|  |  |  |  | t | pFDR | d | t | pFDR | d |
| Left hemisphere | SP-Combined | 0.759<br>(0.006) | 0.755<br>(0.007) | 3.88 | 0.002 | -0.63 | 3.89 | 0.002 | -0.63 |
|  | SP-Position | 0.801<br>(0.005) | 0.797<br>(0.007) | 3.83 | 0.002 | -0.62 | 3.86 | 0.002 | -0.62 |
| Left frontal | SP-Position | 0.807<br>(0.007) | 0.802<br>(0.007) | 3.71 | 0.002 | -0.60 | 3.59 | 0.004 | -0.58 |
| Left temporal | SP-Position | 0.807<br>(0.011) | 0.797<br>(0.011) | 5.21 | < 0.001 | -0.84 | 5.23 | < 0.001 | -0.84 |
|  | SP-Depth | 0.821<br>(0.001) | 0.816<br>(0.011) | 3.02 | 0.019 | -0.49 | 3.17 | 0.010 | -0.51 |
| Right hemisphere | SP-Combined | 0.761<br>(0.006) | 0.758<br>(0.007) | 2.92 | 0.023 | -0.47 | 2.76 | 0.034 | -0.45 |
|  | SP-Topology | 0.896<br>(0.005) | 0.891<br>(0.006) | 4.61 | < 0.001 | -0.74 | 4.55 | < 0.001 | -0.73 |
| Right temporal | SP-SAbasins | 0.879<br>(0.015) | 0.871<br>(0.016) | 3.35 | 0.007 | -0.54 | 3.39 | 0.006 | -0.55 |
| Right parietal | SP-Combined | 0.746<br>(0.012) | 0.741<br>(0.011) | 2.68 | 0.041 | -0.43 | 2.73 | 0.030 | -0.44 |
|  | SP-Topology | 0.891<br>(0.012) | 0.876<br>(0.013) | 7.32 | < 0.001 | -1.18 | 7.36 | < 0.001 | -1.19 |

Details for all comparison results are provided in Supplementary Materials, Tables S2 – S7.

### 3.2. Analysis of the sulcal pattern divergence between groups

When assessing differences in the sulcal patterns between groups (A vs C) in the sex-pooled sample, patients showed a less similar sulcal pits pattern than HC in the left frontal lobe for the SP-Position (mean (SD) HC = 0.806 (0.006); mean (SD) SZ = 0.803 (0.008); t = 3.45; p_FDR_ = 0.03; d = -0.34), although differences did not remain significant after correcting for age (t = 2.79; p_FDR_ = 0.053) (Figure 4A).

**Figure 4.**
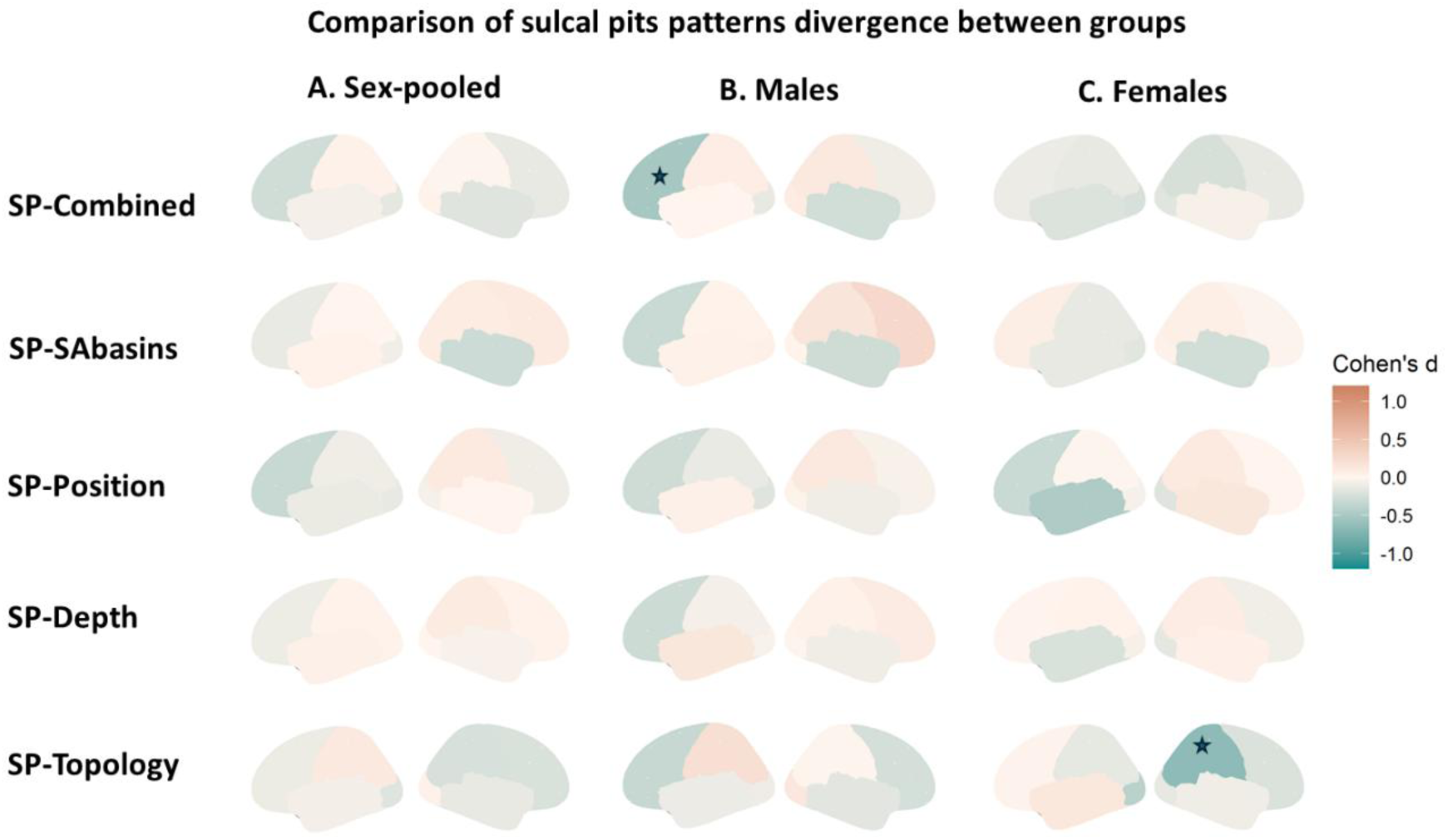
Between group differences in the mean similarity of the sulcal pits patterns. The figure is based on the effect sizes (Cohen’s d) of the differences in sulcal patterns between SZ patients and HC across lobes in the sex-pooled (A), males (B) and females (C) samples. Stars indicate lobes showing age-corrected significant differences.

When performing the analyses separately by sex, the same pattern as in the sex-pooled sample emerged for the SP-Combined in the left frontal lobe, both before adjusting by age (mean (SD) HC = 0.772 (0.008); mean (SD) SZ = 0.767 (0.008); t = 4.22; p_FDR_ = 0.002; d = -0.54) and after (t = 3.98; p_FDR_ = 0.004; d = -0.51) (Figure 4B). On the other hand, females displayed a less similar sulcal pattern than HC as regards SP-Topology of the right parietal lobe (mean (SD) HC = 0.891 (0.012); mean (SD) SZ = 0.882 (0.013); t = 3.92; p_FDR_ = 0.006; d = -0.63) and such differences also remained after correcting by age (4.03; p_FDR_ = 0.004; d = -0.65) (Figure 4C).

Details for all comparison results are provided in Supplementary Materials, Tables S8 – S13.

### 3.3. Association of sulcal patterns with schizophrenia symptomatology

When examining the correlation between sulcal pattern similarity and PANSS scores in those features with significant differences between groups, we observed nominal significant associations exclusively in the male sample. Specifically, we found associations between the left frontal Age-uncorrected SP-Combined and both PANSS total (r = -0.16; p_nom_ = 0.043) and PANSS-GP (r = -0.19; p_nom_ = 0.02). These results indicated that patients with more severe symptoms tended to show sulcal patterns less similar to HC. The results remained nominally significant when using Age-corrected sulcal pattern similarity values for the association between SP-Combined and PANSS-GP (r = -0.16; p_nom_ = 0.04), but not with PANSS total (p_nom_ > 0.05). We did not observe any significant correlation between the right parietal SP-Topology and PANSS in the females sample (p_nom_ > 0.05).

### 3.4. Transcription-neuroimaging association analysis

When conducting the transcription-neuroimaging association analysis, the PLS-R models in males reported that the first component explained 59.2% of the variance in gene expression (X) and 35.4% of the variance in sulcal pattern differences (Y), whereas the second component accounted for 21.8% of the variance in X and 63.2% of the variance in Y. Although the analysis showed a low overall predictability of the model (Q^2^ < 0), the per-variable estimate of performance identified the SP-Topology as the one showing a PRESS/SS > 0.90 (PRESS/SS = 0.66; p = 0.04), making it the most explained variable by the PLS-R model, with R^2^ = 43.22% (Component 1) (Supplementary Material, Table S14). Genes within the top 10% of VIP scores (Supplementary Material, Table S16) were significantly enriched in the following gene ontology (GO) biological processes: 7.82-fold enrichment in the regulation of sodium ion transport (p_FDR_ = 0.036) and 1.85-fold enrichment in nervous system development (p_FDR_ = 0.027) (Supplementary Material, Table S18).

In females, the first component explained 46.8% of the variance in X and 40.7% of the variance in Y, while the second component explained 29.5% of X and 35.3% of Y. SP-Depth emerged as the most explained Y-variable, with R^2^ = 50.6% (Component 1) (Supplementary Material, Table S15), although the overall predictability of the model was low (Q^2^ < 0) and no variable showed a PRESS/SS < 0.90) (Supplementary Material, Table S15), indicating a lower predictive performance of the model for all the Y-variables in females. Among the genes with the top 10% VIP scores (Supplementary Material, Table S17), significant enrichment was observed in central nervous system development (2.40-fold, p_FDR_ = 0.038) (Supplementary Material, Table S19).

## 4. DISCUSSION

This study identified sulcal pits as early developmental markers of neuroanatomical differences between individuals with SZ and HC, particularly in terms of increased heterogeneity and divergence from a healthy control-based average. In addition, our study, the first to analyze the patterns of sulcal pits separately in males and females with schizophrenia, shows that in males, greater divergence in sulcal pits patterns tends to be associated with symptom severity. Also, our results suggest that differences in the heterogeneity of these patterns are linked to specific biological processes, providing some insights into the ontogenetic basis of sulcal pits variability and its relevance to the neurobiology of SZ.

When examining the heterogeneity of sulcal pits patterns (Figure 2, A vs B), patients with SZ consistently exhibited greater heterogeneity compared to HC across most measures and lobes. This is consistent with previous studies reporting increased regional heterogeneity in various neuroanatomical measures among individuals with SZ. For example, greater variability in cortical surface area and cortical thickness have been observed in frontal, temporal, and hippocampal regions (Baldwin et al., 2022). Similarly, a meta-analysis by Brugger & Howes (2017) identified increased volumetric variability in SZ within the temporal lobe, putamen, thalamus, and third ventricle, whereas other regions, such as the cingulate cortex and amygdala exhibited reduced variability compared to HC. The specific regions showing greater heterogeneity in our study do not fully overlap with those identified in previous research. These discrepancies may be attributed to variations in methodological approaches, the choice of brain parcellations, and the specific neuroanatomical phenotype assessed (none of the aforementioned studies were based on sulcal pits). In this regard, it is worth mentioning a study based on sulcal pits graphs comparing the sulcal patterns heterogeneity in individuals with attention deficit/hyperactivity disorder and HC also reported a higher heterogeneity in patients than HC across all lobes (Li, Wang, et al., 2021), aligning with our results. Overall, these findings highlight the relevance of increased heterogeneity in sulcal pits patterns as a marker of neurodevelopmental alterations, suggesting that the brain’s attempt to buffer risk-related inputs during development may lead to heterogeneous pathophysiological trajectories potentially contributing to the outcome and course variability within SZ and other psychiatric disorders.

When examining differences in the divergence of the patterns of sulcal pits between groups (Figure 2, A vs C), no significant differences in sulcal pits pattern divergence were detected in the sex-pooled sample, while they emerged when stratifying the analysis by sex. In males, patients compared to HC showed an averaged reduced similarity in the SP-Combined feature of the left frontal lobe than the averaged similarity within HC. In females, a reduced averaged similarity emerged in patients compared to HC as compared with the averaged similarity within HC in the SP-Topology of the right parietal lobe. Notably, in our results, the regions showing the greatest heterogeneity in sulcal pits patterns among patients were the same regions in which significant differences in the divergence from the averaged HC were observed, highlighting the contribution of patient-specific heterogeneity to the observed group-level divergences.

Despite sulcal pits being considered stable markers (Meng et al., 2014; Solhtalab et al., 2025), recent data have highlighted the importance of considering age effects due to the interaction between age and the number of sulcal pits in a previous study (Hostalet, et al., 2025). Accordingly, we tested whether our results were influenced by age. In the case of sulcal pits pattern heterogeneity, the observed differences in SP-Position and SP-Depth in the right parietal lobe were no longer significant after age adjustment, suggesting that age contributed to the observed heterogeneity in these specific features in the sex-pooled sample. In contrast, regarding the observed differences in the divergence of sulcal pits patterns between groups, none changed when corrected for age. Then, the fact that the effect of age was observed exclusively in certain regions of the right hemisphere in the sex-pooled sample, indicates that such effect could be related to the specific interaction between age and sex on the number of sulcal pits in the right hemisphere reported in healthy individuals in Hostalet et al. (2025). However, a stronger assumption of this fact is not possible, as different sulcal pits assessment methodologies were used in the current study and in Hostalet et al. (2025).

In line with our results highlighting the left frontal lobe as a key region for the divergence in the patterns of sulcal pits between SZ and HC, previous studies have consistently reported structural alterations in the frontal lobe among individuals with SZ linking them to specific SZ clinical and cognitive symptoms (M’Barek et al., 2022; Smucny et al., 2022; Snelleksz et al., 2022). In contrast, alterations in the parietal lobe are less frequently documented, although some evidence points towards a dysconnectivity in this region among individuals with SZ, which may contribute to impairments in multisensory information processing associated with the disorder (Gröhn et al., 2022). The relative scarcity of sex-specific findings, undetectable in our study when males and females were analyzed together, may stem from the common practice of conducting analyses in sex-pooled samples, potentially overlooking sex-specific neuroanatomical patterns associated with the disorder. Additionally, the regions showing the largest effect size differences in sulcal pattern similarity between individuals with attention deficit hyperactivity disorder and HC are consistent with the regions in which we found significant differences in our sample, the left frontal and right parietal lobes (Li, Wang, et al., 2021).

Regarding the association between clinical symptoms and sulcal pits patterns, we observed a trend toward negative correlations between the left frontal SP-Combined feature and both the PANSS PANSS-GP and PANSS scores in the males sample, indicating that patients whose sulcal pits patterns deviated more from the HC average exhibited greater symptom severity. However, these associations did not remain significant after correction for multiple comparisons. This finding aligns with previous research showing that reduced similarity to a normative model of sulcal width based on HC was associated with poorer cognitive performance in individuals with SZ (Janssen et al., 2021). Additionally, previous studies have identified specific patterns of sulcal pits in patients experiencing auditory hallucinations, specifically regarding the frequency of sulcal pits in the temporal lobe (Lerosier et al., 2024) and the depth of sulcal pits in certain regions of the frontal lobe (Salgado-Pineda et al., 2025). Nevertheless, none of these studies specifically examined the association between sulcal pits patterns and symptom severity, as was done in the present study. The absence of a correlation between the right parietal SP-Topology feature and clinical symptoms in females may stem from the smaller sample size of females with SZ compared to males (64 vs. 173), resulting in reduced statistical power, or alternatively from underlying neurobiological sex differences; however, our data do not permit firm conclusions and instead underscore the need for replication in larger samples.

Regarding the correlation between sulcal pits patterns and genetic expression, the low overall predictive performance of the PLS-R models, likely due to the limited number of outcomes variables, indicate that results may be cautiously interpreted and considered as hypothesis-generating rather than predictive. However, in the per-variable predictability assessment in males, SP-Topology emerged as the feature significantly influenced by genetic expression. Genes contributing most strongly to this association between differences in the heterogeneity of sulcal pits and genetic expression, were significantly enriched in biological processes related to sodium ion transport and nervous system development. This finding is aligned with evidence suggesting that SZ is related to neural excitability, synaptic organization and neurotransmission (Trubetskoy et al., 2022). Furthermore, alterations in the sodium channels have been associated with different neurodevelopmental psychiatric disorders (Imbrici et al., 2013).

In females, gene enrichment analysis revealed a significant overrepresentation of genes involved in central nervous system development, suggesting alignment with the neurodevelopmental hypothesis of SZ (Birnbaum & Weinberger, 2024). However, these results did not reach statistical significance, and therefore further analyses in larger samples are needed to explore sex-specific patterns in the genetic contribution to sulcal pits variability. The existence of differential contributions between sexes has been already reported in other neuroanatomical measures such as the local gyrification index, a measure closely related to sulcal pits (Han et al., 2023).

Some limitations should be considered in the present study. First, although the overall sample size is moderate (to our knowledge, the largest to date in studies of sulcal pits patterns in SZ), it becomes considerably smaller when stratified by sex, particularly in the female subgroup. This reduction in sample size may limit the statistical power of our sex-specific analyses. Nevertheless, it is precisely in these sex-stratified analyses that meaningful differences in sulcal pits divergence emerged, highlighting the importance of investigating sex-specific neuroanatomical patterns in SZ.

Regarding the correlation between sulcal pits pattern differences and genetic expression, the main constraint of the applied approach is linked to the consideration of the strengths and limitations of neuroimaging transcriptomics. In this sense, the combination of two datasets, *in vivo* estimates of sulcal pits patterns differences and gene expression data derived from brain postmortem samples, provides a powerful framework to link macroscale imaging phenotypes with underlying molecular mechanisms. However, current transcriptomic resources are derived from a small number of samples susceptible to brain postmortem artifacts and the associations remain correlational, underscoring the need for replication in larger and more diverse datasets as well as integration with longitudinal and cell-type–specific data. Nonetheless, to add robustness to this approach, we restricted our analysis to genes exhibiting stable inter-regional expression profiles across two independent datasets, as explained in Shin et al. (2018).

## 5. CONCLUSION

This study provides novel evidence supporting sulcal pits as early neurodevelopmental markers of structural brain variability in SZ. By examining both the heterogeneity and divergence of sulcal pits patterns, we identified sex-specific alterations marginally linked to symptom severity and distinct biological processes. The identification of sex-specific genetic enrichment patterns underscores the need for more targeted, stratified approaches in psychiatric research. These results not only advance our understanding of the developmental origins of brain structure in SZ but also provide a basis for future research aimed at refining neurobiological subtyping linked to individual variability in clinical presentation.

## Data Availability

All data produced in the present study are available upon reasonable request to the authors.

## AUTHOR CONTRIBUTIONS

Conceptualization: Noemí Hostalet, Kiho Im and Mar Fatjó-Vilas

Data curation: Noemí Hostalet

Formal analysis: Noemí Hostalet, Daniel Herrera-Escartín, Mateo Berrio

Investigation: Noemí Hostalet, Amalia Guerrero-Plaza, Pedro Canut, Jesús J Gomar, Pilar

Salgado-Pineda, Josep Salvaert, Nuria Ramiro, Emili Inarejos-Clemente, Salvador Sarro, Raymond Salvador, Edith Pomarol-Clotet, Mar Fatjó-Vilas.

Methodology: Noemí Hostalet, Lluís Cobos-Aumatell, Banu Ahtam, Edith Pomarol-Clotet, Kiho Im, Mar Fatjó-Vilas

Visualization: Noemí Hostalet; Lluís Cobos-Aumatell

Resources: Edith Pomarol-Clotet and Mar Fatjó-Vilas

Funding acquisition: Neus Martínez-Abadías, Edith Pomarol-Clotet and Mar Fatjó-Vilas

Supervision: Mar Fatjó-Vilas

Writing – original draft: Noemí Hostalet and Mar Fatjó-Vilas

Writing – review & editing: All authors.

## Funding

This study received funding from: i) Instituto de Salud Carlos III (ISCIII) through the project PI20/01002, the contracts FI21/00093 to NH and CP20/00072 to MF-V and the mobility grant MV23/00059 to NH (co-funded by European Regional Development Fund (ERDF)/European Social Fund “Investing in your future”); ii) Agencia Estatal de Investigación through the project PID2020-113609RB-C21 and the grant PRE2021-100310 to DH-E, funded by MICIU/AEI/10.13039/501100011033 and “ESF Investing in your future”; iii) Comissionat per a Universitats i Recerca del DIUE of the Generalitat de Catalunya (Agència de Gestió d’Ajuts Universitaris i de Recerca (AGAUR): 2021SGR01475 and 2021SGR00706).

## Ethical Statement

Ethical approval was obtained from the FIDMAG Hermanas Hospitalarias Research Ethics Committee (protocol number PR-2023-13). All participants provided written informed consent about the study procedures and implications, which were carried out according to the Declaration of Helsinki.

## Availability of data and materials

The data supporting the findings of this study are available from the corresponding authors upon reasonable request.

## Consent for publication

Not applicable

## Competing interests

The authors declare that they have no competing interests.

## Acknowledgements

The authors thank all the participants; without their generosity this study would not have been possible.

## Notes

### Competing Interest Statement

The authors have declared no competing interest.

### Author Declarations

Ethical approval was obtained from the FIDMAG Hermanas Hospitalarias Research Ethics Committee (protocol number PR-2023-13).

